# The association between chrononutrition behaviors and muscle health among older adults: The Study of Muscle, Mobility and Aging (SOMMA)

**DOI:** 10.1101/2023.11.13.23298454

**Authors:** Ziling Mao, Peggy M Cawthon, Stephen B Kritchevsky, Frederico G S Toledo, Karyn A Esser, Melissa L Erickson, Anne B Newman, Samaneh Farsijani

## Abstract

**Background:** Emerging studies highlight chrononutrition’s impact on body composition through circadian clock entrainment, but its effect on older adults’ muscle health remains largely overlooked.

**Objective:** To determine the associations between chrononutrition behaviors and muscle health in older adults.

**Methods:** Dietary data from 828 older adults (76±5y) recorded food/beverage amounts and their clock time over the past 24 hours. Studied chrononutrition behaviors included: **1)** <u>The clock time of the first and last</u> food/beverage intake; **2)** <u>Eating window</u> (the time elapsed between the first and last intake); and **3)** <u>Eating frequency</u> (Number of self-identified eating events logged with changed meal occasion and clock time). Muscle mass (D_3_-creatine), leg muscle volume (MRI), grip strength (hand-held dynamometer), and leg power (Keiser) were used as outcomes. We used linear regression to assess the relationships between chrononutrition and muscle health, adjusting for age, sex, race, marital status, education, study site, self-reported health, energy, protein, fiber intake, weight, height, and moderate-to-vigorous physical activity.

**Results:** Average eating window was 11±2 h/d; first and last intake times were at 8:22 and 19:22, respectively. After multivariable adjustment, a longer eating window and a later last intake time were associated with greater muscle mass (β±SE: 0.18±0.09; 0.27±0.11, respectively, *P*<0.05). The longer eating window was also marginally associated with higher leg power (*P*=0.058). An earlier intake time was associated with higher grip strength (−0.38±0.15; *P*=0.012).

**Conclusions:** Chrononutrition behaviors, including longer eating window, later last intake time, and earlier first intake time were associated with better muscle mass and function in older adults.

**GRAPHICAL ABSTRACT:** 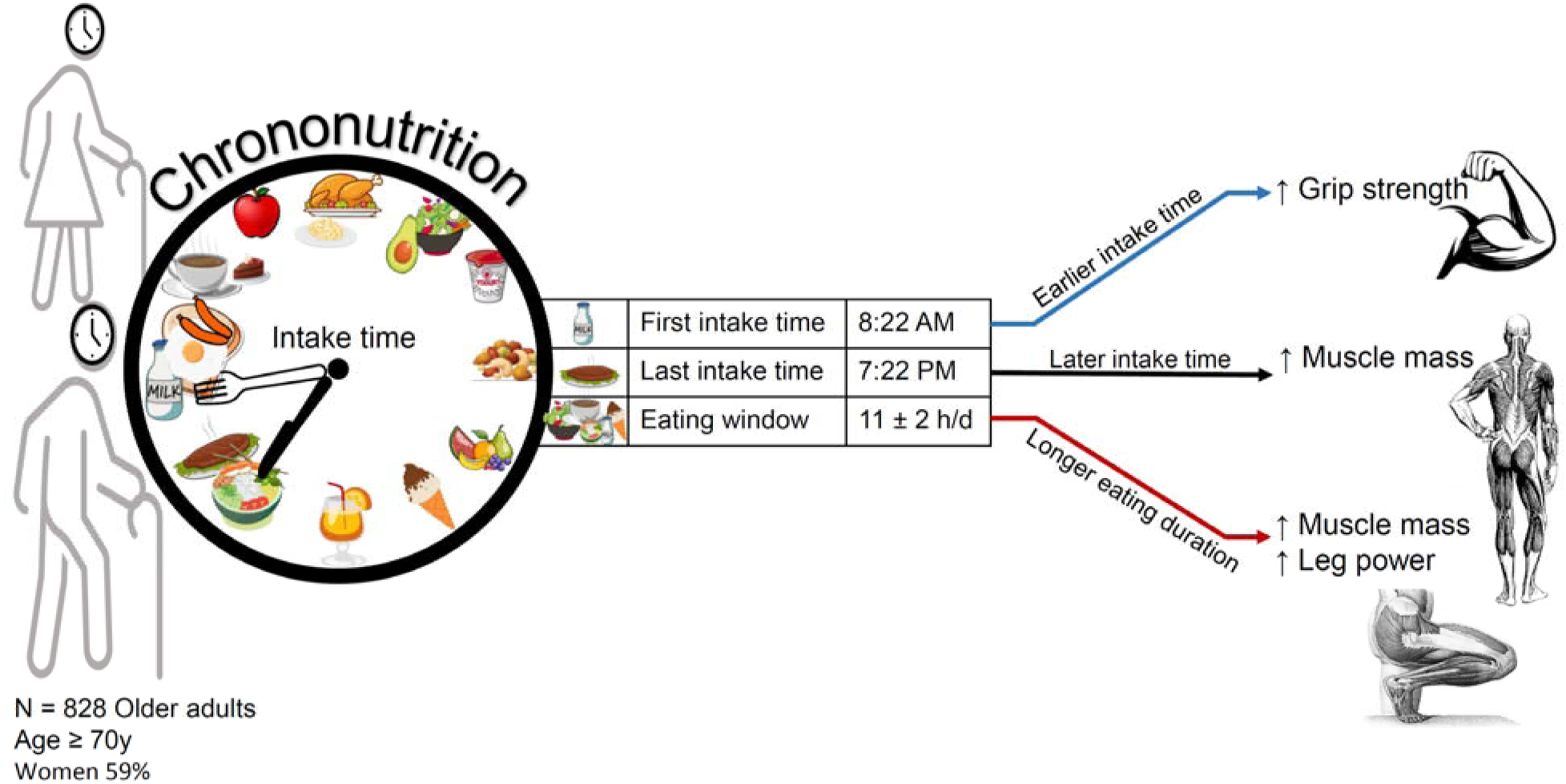

**Key findings:** Chrononutrition behaviors, including longer eating window, later last intake time, and earlier first intake time were associated with better muscle mass and function in older adults.

## INTRODUCTION

Sarcopenia is an aging-related disease characterized by loss of muscle mass and reduced muscle functions, and it is associated with higher risk of disability and mortality among older adults [1, 2]. Several dietary factors, such as protein intake [3, 4], vitamin D supplementation [5], Mediterranean diet [6], and fasting regimens [7] have been suggested to maintain muscle mass and function in older adults. While many studies have focused on the quality and quantity of dietary intake to promote muscle health in aging, the relationship between the timing and frequency of food and beverage intake, also known as chrononutrition [8], and muscle health remains unclear.

A growing body of research has revealed that the timing of food/beverage consumption can impact body composition and overall health by entraining the body’s circadian clocks, i.e., peripheral clocks located in various organs, including muscles [8, 9]. It has been shown that inappropriate meal timing patterns, such as skipping breakfast or eating late at night, can disrupt the peripheral tissue clocks [10]. This circadian disruption may have adverse effects on metabolic processes, leading to disturbances in lipid and glucose metabolism [11, 12], insulin resistance [13], and ultimately, changes in body composition, including alterations in body weight and fat mass [14–18]. However, the importance of chrononutrition on muscle mass and function remains unknown. The timing of food and beverage consumption is also an external cue [9] that can modulate the expression of clock genes located in skeletal muscle that play a crucial role in the circadian regulation of skeletal muscle physiology and maintaining optimal muscle structure and function [19]. Small-scale human studies on healthy young adults have shown that breakfast skipping [16, 20, 21] or time-restricted eating [17] were associated with lower muscle mass. In recent years, various types of intermittent fasting diets, particularly time-restricted feeding, which involves reducing the daily eating window, have been proposed to offer multiple health benefits [22]. Nevertheless, a significant proportion of the research on intermittent fasting diets has been performed on animal models [23] or young and physically active adults [24, 25]. Therefore, there is a need for large-scale human studies to determine the role of chrononutrition in maintaining muscle health in older adults.

Our group previously showed that an even distribution pattern of protein intake throughout the day, with equal amounts at breakfast, lunch, and dinner, was significantly associated with higher muscle mass [26] and greater physical function [27] in older adults compared to a skewed pattern of protein consumption at one meal, usually at dinner, regardless of the total amount of protein consumed. However comprehensive assessments of other chrononutrition behaviors are needed to better understand how timing of food/beverage intake contributes to muscle health beyond conventional nutritional factors. Ultimately, this will lead to the development of precision nutrition strategies for older adults that can be tested in clinical trials, by guiding what and when to eat in order to meet nutrient needs, prevent sarcopenia, and promote health.

Therefore, our study aimed to determine the cross-sectional association between multiple chrononutrition behaviors and muscle health among older adults (≥70 years old) from the Study of Muscle, Mobility and Aging (SOMMA). These chrononutrition behaviors included the timing of the first and last food/beverage intake, eating window, and eating frequency. Our study outcomes included measures of muscle mass, thigh muscle volume, grip strength, and leg power.

## METHODS

### Study population

SOMMA is a longitudinal study of mobility and muscle among community-dwelling older adults conducted at two clinical sites: University of Pittsburgh (Pittsburgh, PA) and Wake Forest University School of Medicine (Winston-Salem, NC) [28]. Details on SOMMA recruitment and study protocols were previously reported [28]. In brief, participants aged 70 years or older, with a body mass index (BMI) of 18-40 kg/m^2^, who were able to complete a muscle tissue biopsy and magnetic resonance (MR) imaging, walk a quarter mile, climb a flight of stairs, and who had no active cancer or advanced chronic diseases (i.e., heart failure, dementia, renal failure on dialysis, or Parkinson’s Disease) were eligible for inclusion. At baseline, SOMMA participants underwent *three* separate clinic visits within a 4–6-week window during which a comprehensive set of assessments was conducted. These assessments included collection of biospecimens, such as muscle and adipose tissue, blood, urine, and fecal samples; completion of various questionnaires, including those related to diet; physical and cognitive function evaluations; whole-body imaging; and cardiopulmonary exercise testing. The study protocol was approved by the Western Institutional Review Board Copernicus Group (WCG IRB; study number 20180764), and written informed consent was acquired from all participants.

Participants in the present study were from SOMMA baseline visits, enrolled between April 2019 and December 2021. Of 879 participants who completed baseline visit across two clinical sites, we excluded those with missing and implausible dietary data (e.g., missing food records or incomplete 24-h food recall; n=51). Our final analytical sample included 828 individuals (**Supplemental Figure 1**).

### Dietary assessment

An online public-use Automated self-administered 24-hour Food Recall (ASA24) developed by the National Cancer Institute (NCI) [29] was used to collect detailed information on the types and amounts of foods and beverages consumed as well as their clock time over the past 24-hour period (from 12:00 AM to 11:59 PM). At the screening visit, each participant was given a worksheet to complete at home, which helped to jog their memory on all food/beverage intakes for a specific day and a research staff member assisted participants in reporting their dietary intake and entering it into the ASA24 website. Two ASA24 food recalls were collected per participant, on two out of the three separate baseline clinic visits. Among participants who recorded their food intake, 90.9% had two 24-hour food recalls while 9.1% had only one recall. For participants with two ASA24 food recalls, we followed a widely accepted approach [30] by calculating the average value of food/beverage consumed and the time of intake. This approach helped us obtain a more accurate representation of participants’ usual dietary patterns.

ASA24 website uses the reported food/beverage items and their corresponding amounts to estimate the nutrient composition of the reported diet using the USDA National Nutrient Database for Standard Reference and other sources [31]. We calculated total daily energy intake (kcal/day) as the sum of calorie intake from all food and beverage within 24 hours. Daily intake of dietary protein, carbohydrate, and fat was determined based on the proportion of total energy intake from each macronutrient (% of kcal/day). We also determined grams of protein intake per kilogram of the body weight (g/kg BW/day) and amount of daily fiber intake (g/day). The dietary quality of participants was estimated using Healthy Eating Index (HEI) 2015 [32], which is designed by USDA to evaluate adherence to 2015-2020 Dietary Guidelines for Americans. To calculate HEI, we extracted the intake of 13 components from the collected food recalls. These comments included total vegetables, greens and beans, total fruits, whole fruits, whole grains, dairy, total protein foods, seafood and plant proteins and fatty acids, sodium, refined grains, saturated fats and added sugars. The scores for each component were then summed to obtain the HEI score which ranges from 0 to 100, with a higher score indicating a healthier diet. The HEI score was calculated per day for each participant, and the average value of the HEI from the two collected 24-hour food recalls per participant was used for analysis.

### Assessment of chrononutrition behaviors

During the dietary recall interview, participants reported the clock time at which they consumed each food or beverage item for the preceding day. They also self-identified the eating occasion for each item as breakfast, lunch, dinner, snack, or drink. The time at which the first food or beverage containing more than 0 kcal was consumed after midnight (00:00) within a day was defined as the time of the *first* food/beverage consumption (in hours per day) [33]. The clock time of the *last* consumption of food or beverage containing more than 0 kcal before midnight (23:59) within a day was recorded as the designated time for the end of food and beverage consumption (in hours per day) [33]. *Eating window* (in hours per day) was determined by calculating the time between the first and last consumption of food or beverage [33, 34].

Furthermore, we extracted the reported self-identified eating events (i.e., breakfast, lunch, dinner, and snacks) of the participants and their corresponding clock times. Then, we determined the eating frequency (number per day), by counting the total number of self-identified eating events, considering a new eating event whenever both the clock time and the self-identified meal changed [33, 35]. Moreover, we analyzed the proportion of total energy intake attributed to each eating event in order to identify which eating event had the highest or lowest contribution to the total daily energy intake.

### Assessment of muscle mass and thigh muscle volume

*Total muscle mass* (kg) was determined using the D_3_-creatine (D_3_Cr) dilution method [36]. Participants ingested a 30-mg dose of stable isotope labeled creatine, and provided a fasting, morning urine sample 72 - 144 hours later. High-performance liquid chromatography (HPLC) and tandem mass spectrometry (MS) were used to measure D_3_-creatinine, unlabeled creatinine, and creatine. An algorithm was used to determine total body creatine pool size and thus skeletal muscle mass (kg) [37, 38]. Of note, this method is not dependent on creatinine clearance or renal function and does not require special dietary control except for the fasting morning urine sample.

*Thigh muscle volumes* (L) were determined based on MR images which were analyzed using AMRA Researcher® (AMRA Medical AB, Linköping Sweden). Briefly, the image analysis included image calibration [39], fusion of image stacks, image segmentation [40], and quantification of fat and muscle volumes [41–43]. The manual quality control was conducted by a blinded trained operator. Fat-tissue free muscle volume in the thighs, i.e., “viable muscle tissue”, was measured as the volume of all voxels with a fat fraction below 50%.

### Assessment of grip strength and leg power

*Grip strength* (kg) was assessed by a hydraulic, isometric, hand-held dynamometer (Jamar) [44]. Individuals who had hand or wrist surgery (i.e., fusion, tendon repair, arthroplasty, or synovectomy) within the past 12 weeks were excluded from the examination. To ensure proper grip size adjustment, each participant performed one submaximal practice trial before the formal examination. Four formal trials were then conducted (two for each hand) with a rest period of at least 15 to 20 seconds between trials. Participants were instructed to squeeze the handgrip with maximum force, and the kilograms of force were recorded from the instrument. The maximum grip strength was determined as the highest kilograms recorded from any of the four trials.

*Leg power* was assessed by the Keiser Air 420 exercise machine [45]. The dominant leg, unless contraindicated by previous joint replacement or limited range of motion, was selected for the test. Individuals with recent occurrences of stroke, aneurysm, cerebral hemorrhage, or systolic blood pressure outside the range of 90-180 mmHg were excluded from participating. The 1 repetition maximum (1-RM) test commenced with a starting resistance of 40 pounds (the lowest setting) and continued until participants reported inability to proceed with higher resistance. Peak leg power of each trial conducted at 40%, 50%, 60%, and 70% 1-RM were then recorded, and highest peak power was identified as the maximum power achieved in any of the trials. The peak leg power standardized for body weight (Watts/kg), was calculated as the highest peak power divided by the participant’s body weight for all individuals.

### Assessment of covariates

Self-reported demographics, including age, sex (men or women), race (white, black, or others/multiracial), education level (≤ high school and others, some college, college degree, or some graduate/graduate degree), marital status (married, widowed, separated/divorced, or single/never married), health status, and use of medications were collected via questionnaires. We used the validated Community Healthy Activities Model Program for Seniors (CHAMPS) 40-item questionnaire [46, 47] to assess self-reported frequency and duration, and time spent on a wide range of physical activities. The CHAMPS questionnaire inquiries about activities carried out in the past 4 weeks, spanning a spectrum from sedentary and light activities (e.g., reading, attending concerts, leisurely walking, and household chores) to moderate-to-vigorous physical activities (MVPA), including jogging, brisk walking, playing basketball, or engaging in moderate to heavy strength training). Each activity was categorized into various levels of physical activity intensity based on metabolic equivalents (METs) [47]. We used the following derived summary metrics from the CHAMPS in our statistical analysis: the sum of total time spent (hours per week) on all exercise-related physical activities (METs ≥ 2) and moderate-to-vigorous physical activity (MVPA, METs ≥ 3). Self-rated health was assessed using a 5-point Likert scale (excellent, very good, good, fair, and poor). Measured height (meter; stadiometer) and body weight (kg; balance beam scale) were used to calculate body mass index (BMI, kg/m^2^). The 10-item screening questionnaire of Center for Epidemiologic Studies Depression Scale (CESD-10) was used to assess depression symptoms [48].

### Statistical analyses

Descriptive analyses were used to summarize the participant characteristics according to the tertiles of eating window. The differences across the tertiles were assessed using chi-square tests for categorical variables, one-way ANOVA for normally distributed, and Kruskal-Wallis non-parametric test for non-normally distributed continuous variables.

General linear regression models were used to assess the associations between the chrononutrition variables (i.e., eating window, first and last time of food/beverage intake, and eating frequency) and muscle outcomes (i.e., muscle mass, thigh muscle volume, grip strength, and leg power). We fitted an unadjusted model and a multivariable linear model which adjusted for the following variables associated with both independent and dependent variables based on our correlation analyses: age (years), sex (men or women), race (white, black, or others/multiracial), total daily energy (kcal/d), protein (%kcal/d), and fiber (g/d) intake, weight (kg), height (meters), MVPA (hours/week), site (Pittsburgh or Wake Forest), education level (≤ high school and others, some college, college degree, or some graduate/graduate degree), marital status (married, widowed, separated/divorced, or single or never married), and self-reported health status (excellent, very good, good, fair, or poor). Of note, because the interaction terms between study predictors and sex were not statistically significant, we opted not to perform regression models stratified by sex. All analyses were completed in SAS version 9.40 (SAS Institute, Cary, NC). Two-sided P-values < 0.05 or 95% confidence intervals (95% CI) not including 0 were considered as statistically significant.

## RESULTS

Out of 828 participants, 59% were women, 13% were black, and the mean age of our study participants was 76 ± 5 years (**Table 1**). Most participants had a college or post-college level of education, were married, and in very good health. The majority of them were enrolled after the Covid-19 Pandemic began (March 2020). The mean BMI of participants was 27.6 kg/m^2^.

**Table 1.**
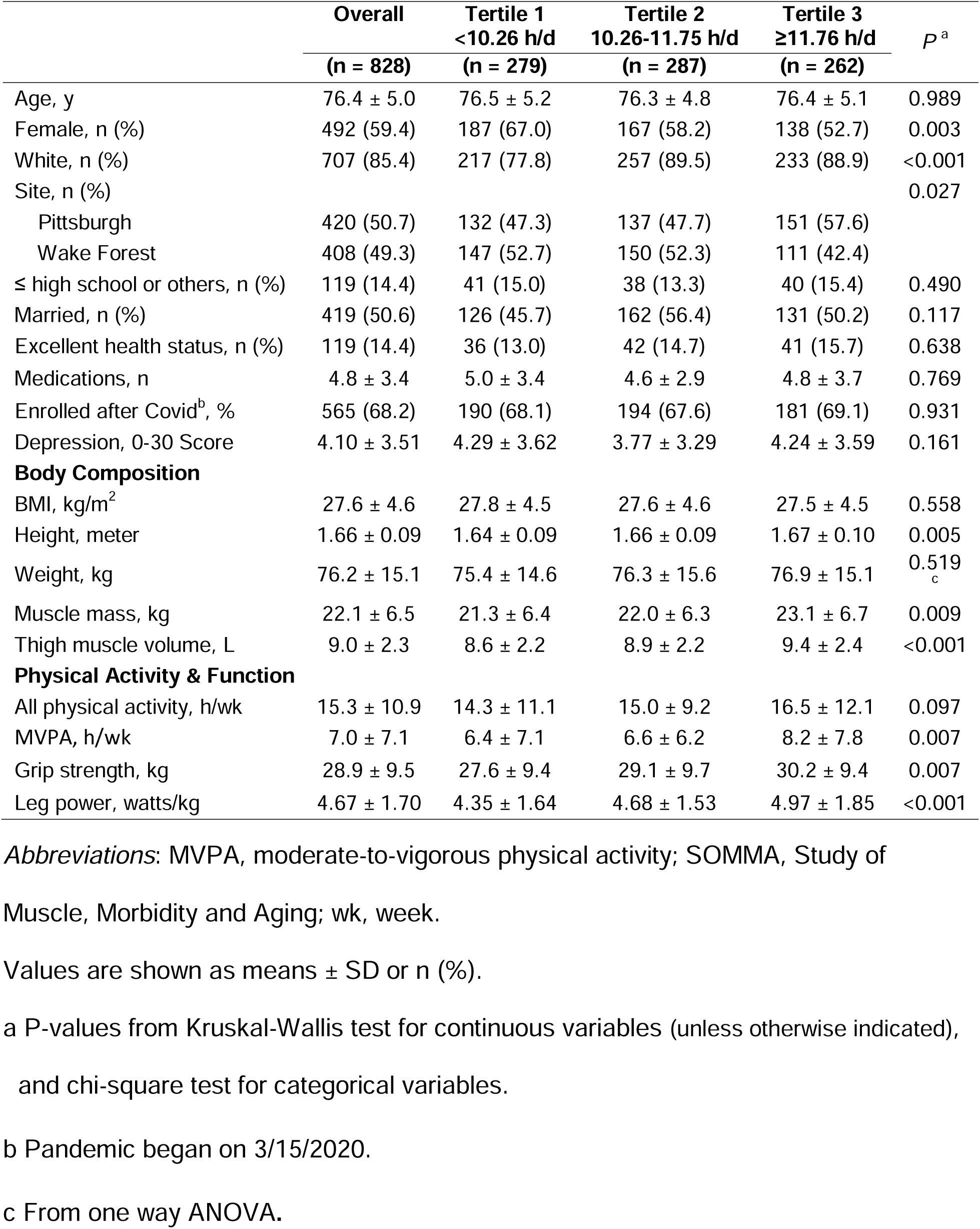
Characteristic of 828 participants in the SOMMA study, presented overall and according to tertiles of eating window.

|  | **Overall** | **Tertile 1** | **Tertile 2** | **Tertile 3** | *P*<sup>a</sup> |
| --- | --- | --- | --- | --- | --- |
|  | **(n = 828)** | **<10.26 h/d**<br>**(n = 279)** | **10.26-11.75 h/d**<br>**(n = 287)** | **≥11.76 h/d**<br>**(n = 262)** |  |
| Age, y | 76.4 ± 5.0 | 76.5 ± 5.2 | 76.3 ± 4.8 | 76.4 ± 5.1 | 0.989 |
| Female, n (%) | 492 (59.4) | 187 (67.0) | 167 (58.2) | 138 (52.7) | 0.003 |
| White, n (%) | 707 (85.4) | 217 (77.8) | 257 (89.5) | 233 (88.9) | <0.001 |
| Site, n (%) |  |  |  |  | 0.027 |
| Pittsburgh | 420 (50.7) | 132 (47.3) | 137 (47.7) | 151 (57.6) |  |
| Wake Forest | 408 (49.3) | 147 (52.7) | 150 (52.3) | 111 (42.4) |  |
| ≤ high school or others, n (%) | 119 (14.4) | 41 (15.0) | 38 (13.3) | 40 (15.4) | 0.490 |
| Married, n (%) | 419 (50.6) | 126 (45.7) | 162 (56.4) | 131 (50.2) | 0.117 |
| Excellent health status, n (%) | 119 (14.4) | 36 (13.0) | 42 (14.7) | 41 (15.7) | 0.638 |
| Medications, n | 4.8 ± 3.4 | 5.0 ± 3.4 | 4.6 ± 2.9 | 4.8 ± 3.7 | 0.769 |
| Enrolled after Covid<sup>b</sup>, % | 565 (68.2) | 190 (68.1) | 194 (67.6) | 181 (69.1) | 0.931 |
| Depression, 0-30 Score | 4.10 ± 3.51 | 4.29 ± 3.62 | 3.77 ± 3.29 | 4.24 ± 3.59 | 0.161 |
| **Body Composition** |  |  |  |  |  |
| BMI, kg/m<sup>2</sup> | 27.6 ± 4.6 | 27.8 ± 4.5 | 27.6 ± 4.6 | 27.5 ± 4.5 | 0.558 |
| Height, meter | 1.66 ± 0.09 | 1.64 ± 0.09 | 1.66 ± 0.09 | 1.67 ± 0.10 | 0.005 |
| Weight, kg | 76.2 ± 15.1 | 75.4 ± 14.6 | 76.3 ± 15.6 | 76.9 ± 15.1 | 0.519<sub>c</sub> |
| Muscle mass, kg | 22.1 ± 6.5 | 21.3 ± 6.4 | 22.0 ± 6.3 | 23.1 ± 6.7 | 0.009 |
| Thigh muscle volume, L | 9.0 ± 2.3 | 8.6 ± 2.2 | 8.9 ± 2.2 | 9.4 ± 2.4 | <0.001 |
| **Physical Activity & Function** |  |  |  |  |  |
| All physical activity, h/wk | 15.3 ± 10.9 | 14.3 ± 11.1 | 15.0 ± 9.2 | 16.5 ± 12.1 | 0.097 |
| MVPA, h/wk | 7.0 ± 7.1 | 6.4 ± 7.1 | 6.6 ± 6.2 | 8.2 ± 7.8 | 0.007 |
| Grip strength, kg | 28.9 ± 9.5 | 27.6 ± 9.4 | 29.1 ± 9.7 | 30.2 ± 9.4 | 0.007 |
| Leg power, watts/kg | 4.67 ± 1.70 | 4.35 ± 1.64 | 4.68 ± 1.53 | 4.97 ± 1.85 | <0.001 |

*Abbreviations:* MVPA, moderate-to-vigorous physical activity; SOMMA, Study of Muscle, Morbidity and Aging; wk, week.
Values are shown as means ± SD or n (%).
a P-values from Kruskal-Wallis test for continuous variables (unless otherwise indicated), and chi-square test for categorical variables.
b Pandemic began on 3/15/2020.
c From one way ANOVA.

### Nutrient intakes and chrononutrition behaviors of SOMMA participants

Our study population had a mean daily eating window of 11.0 hours, with the first and last food/beverage intake occurring at a mean clock time of 8:22 and 19:22, respectively. The participants had a mean meal frequency of 4.1 times per day (**Table 2**). Women had a shorter mean daily eating window than men, 10.8 hours/day compared to 11.3 hours/day, respectively (*P* < 0.01; **Figure 1**). The eating window for white participants was 11.1 hours/day, whereas black participants had a shorter duration at 10.3 hours/day (*P* < 0.01) and other/multiracial participants had an even shorter duration at 9.9 hours/day (*P* < 0.01) (**Figure 1**). Women had a slightly later average breakfast intake time than men: 8.6 *vs* 8.3 hours/day (*P* < 0.01). Among all racial groups, black participants had the latest breakfast (9.1 hours/day) and lunch (13.5 hours/day) times. This compares to 8.4 hours/day for breakfast and 12.9 hours/day for lunch for white participants, and 8.4 and 12.6 hours/day for other/multiracial participants, respectively (*P* < 0.01) (**Figure 1**).

**Figure 1.**
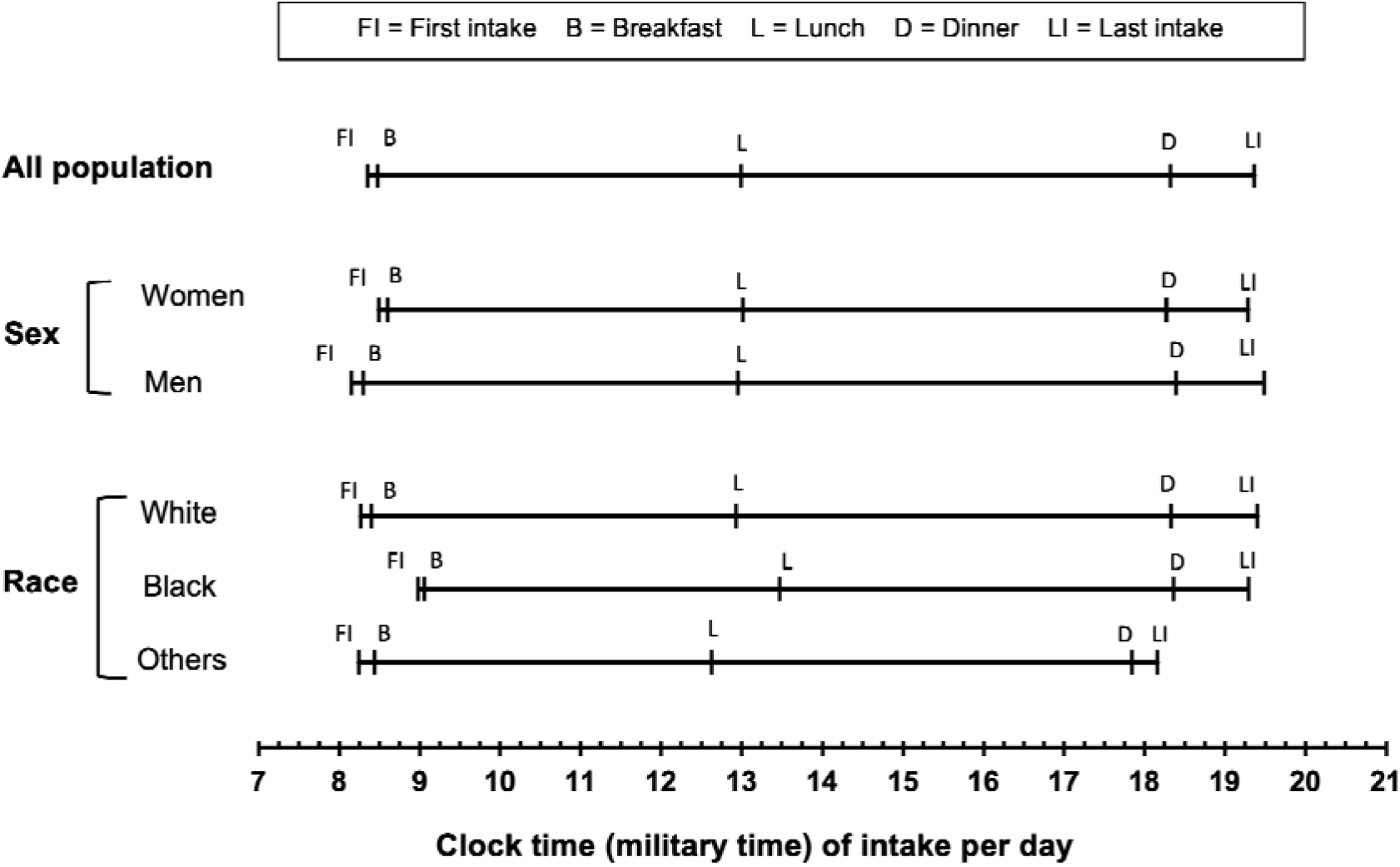
Mean clock time of different chrononutrition behaviors among SOMMA participants (N = 828), overall and across different population subgroups. Significant differences were observed in the time of first food/beverage consumption (*P* = 0.001) and breakfast time (*P* = 0.002) between men and women. Also, across race groups, there were significant differences in the first (*P* < 0.001), last (*P* = 0.002) intake time as well as the intake time of breakfast (*P* < 0.001) and lunch (*P* < 0.001).

**Table 2.** Nutrient intakes and chrononutrition behaviors among SOMMA participants, presented overall and according to tertiles of eating window.

|  | Overall | Tertile 1<br><10.26 h/d | Tertile 2<br>10.26-11.75 h/d | Tertile 3<br>≥11.76 h/d | P<sup>a</sup> |
| --- | --- | --- | --- | --- | --- |
|  | (n = 828) | (n = 279) | (n = 287) | (n = 262) |  |
| **Dietary intakes** |  |  |  |  |  |
| Energy intake, kcal/d | 1678 ± 493 | 1556 ± 470 | 1668 ± 467 | 1819 ± 510 | <0.001 |
| Protein, %kcal/d | 16.9 ± 4.3 | 17.1 ± 4.5 | 17.1 ± 4.4 | 16.5 ± 4.0 | 0.224 |
| Carbohydrates, %kcal/d | 47.0 ± 8.9 | 46.3 ± 9.0 | 47.2 ± 9.2 | 47.5 ± 8.3 | 0.264<sup>b</sup> |
| Fat, %kcal/d | 36.0 ± 7.1 | 36.7 ± 7.5 | 35.7 ± 7.1 | 35.6 ± 6.8 | 0.135<sup>b</sup> |
| Protein, g/kg/d | 0.94 ± 0.35 | 0.88 ± 0.32 | 0.94 ± 0.34 | 0.98 ± 0.37 | 0.002 |
| Fiber, g/d | 17.5 ± 7.8 | 16.7 ± 7.8 | 17.4 ± 7.2 | 18.6 ± 8.3 | 0.005 |
| HEI, scores 0-100 | 58.4 ± 11.8 | 57.7 ± 11.4 | 59.2 ± 12.0 | 58.2 ± 12.1 | 0.274<sup>b</sup> |
| **Chrononutrition variables** |  |  |  |  |  |
| Time of first intake, h/d | 8.35 ± 1.49 | 9.29 ± 1.38 | 8.30 ± 1.06 | 7.40 ± 1.37 | <0.001 |
| Time of last intake, h/d | 19.37 ± 1.48 | 18.30 ± 1.24 | 19.35 ± 1.08 | 20.50 ± 1.20 | <0.001 |
| Eating window, h/d | 11.01 ± 1.92 | 9.01 ± 1.21 | 11.04 ± 0.42 | 13.10 ± 1.11 | <0.001 |
| Eating frequency, n/d | 4.14 ± 0.95 | 3.59 ± 0.80 | 4.18 ± 0.82 | 4.66 ± 0.88 | <0.001 |
| Breakfast, %kcal/d | 24.0 ± 11.8 | 25.0 ± 13.2 | 24.5 ± 11.3 | 22.3 ± 10.6 | 0.014 |
| Lunch, %kcal/d | 26.1 ± 13.1 | 25.0 ± 14.9 | 26.7 ± 12.4 | 26.4 ± 11.7 | 0.261<sup>b</sup> |
| Dinner, %kcal/d | 35.3 ± 13.9 | 38.8 ± 15.3 | 34.4 ± 13.2 | 32.6 ± 12.1 | <0.001<sup>b</sup> |
| Snacks, %kcal/d | 14.6 ± 11.9 | 11.1 ± 11.7 | 14.4 ± 10.6 | 18.6 ± 12.4 | <0.001 |
| Weekday recalls, n (%) | 818 (98.8) | 274 (98.2) | 284 (99.0) | 260 (99.2) | 0.523 |

*Abbreviations:* d, day; HEI, healthy eating index (2015); SOMMA, Study of Muscle, Morbidity and Aging.
Values are shown as means ± SD or n (%).
<sup>a</sup> P-values from Kruskal-Wallis test for continuous variables (unless otherwise indicated), and chi-square test for categorical variables.
<sup>b</sup> From one way ANOVA.

Among our study participants dinner attributed to the greatest amount of total daily energy intake (35% of total daily energy intake), followed by lunch (26%) and breakfast (24%), while snacks (15%) had the lowest contributions to total daily energy intake (**Table 2** and **Figure 2**). Energy intake patterns were similar among men and women, however there was a slight difference across racial groups (**Figure 2**). We observed that the proportion of daily energy intake from snacks was higher in black participants than other subgroups (*P* = 0.017). Almost all participants had their food recalls collected on weekdays with only 1% collected on weekends.

**Figure 2.**
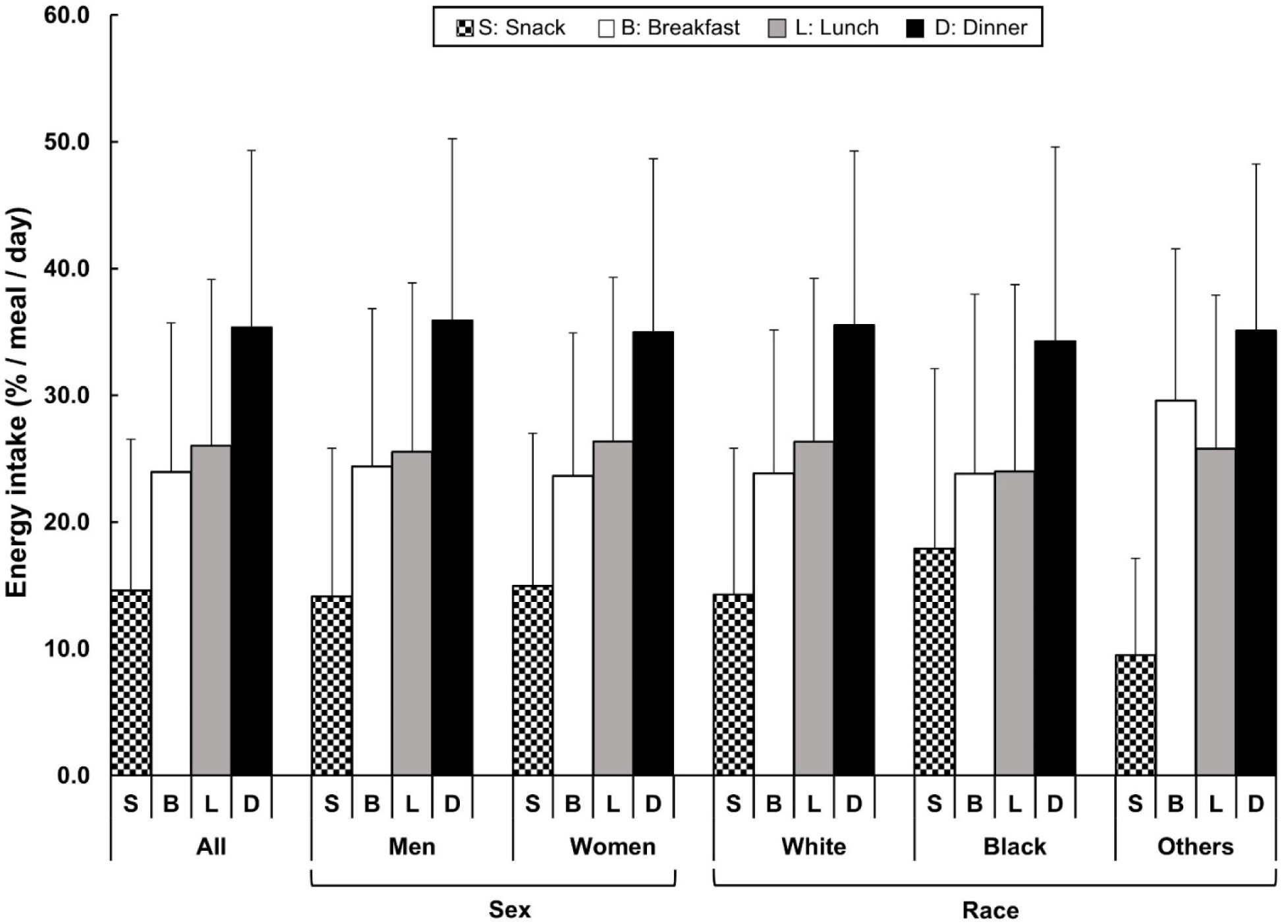
Proportion of energy intake among SOMMA participants (N = 828), based on self-identified eating occasions, including breakfast, lunch, dinner, and snacks. Values are presented as mean ± standard deviation.

### Participant characteristics by eating window tertiles

The participants in the higher tertiles of eating window were predominantly men, white, and from Pittsburgh (**Table 1**). Individuals in both the higher and lower tertiles of eating window were similar in terms of marital status, education, and self-rated health status. Compared to participants in the lower tertiles of eating window, those in the higher tertiles had higher muscle mass, greater thigh muscle volume, grip strength, and leg power, and were also more physically active.

Moreover, participants in the upper tertiles of eating window tended to start eating earlier and finish later, and exhibited higher eating frequency, as expected (**Table 2**). Participants with a longer eating window consumed a significantly greater proportion of their daily energy intake through snacks in comparison to individuals with a shorter eating window. Regarding energy and nutrient intake, individuals in the upper tertiles of eating window exhibited a higher total daily energy intake than those in the lower tertiles, with a similar proportion of calorie intake from each macronutrient across groups (**Table 2**).

### Association between chrononutrition behaviors and muscle mass and volume

We performed linear regression analysis to examine the associations between chrononutrition behaviors and muscle mass and volume. In our unadjusted model, we observed a significant positive association between eating window (h/d) and muscle mass (kg), with a value of β ± standard error (SE) of 0.43 ± 0.12 (**Table 3**). This positive association remained significant after adjusting for age, sex, race, total energy, protein, and fiber intake, weight, height, MVPA, study site, education level, marital status, and self-reported health status (**Table 3** and **Figure 3**). That is, on average, each one-hour increment in daily eating window was associated with 0.18 ± 0.09 kg greater muscle mass (*P* = 0.034). Similarly, we observed a significant positive association between eating window and thigh muscle volume in our unadjusted regression model (**Table 3**), however, this association did not remain significant after adjusting for abovementioned covariates.

**Figure 3.**
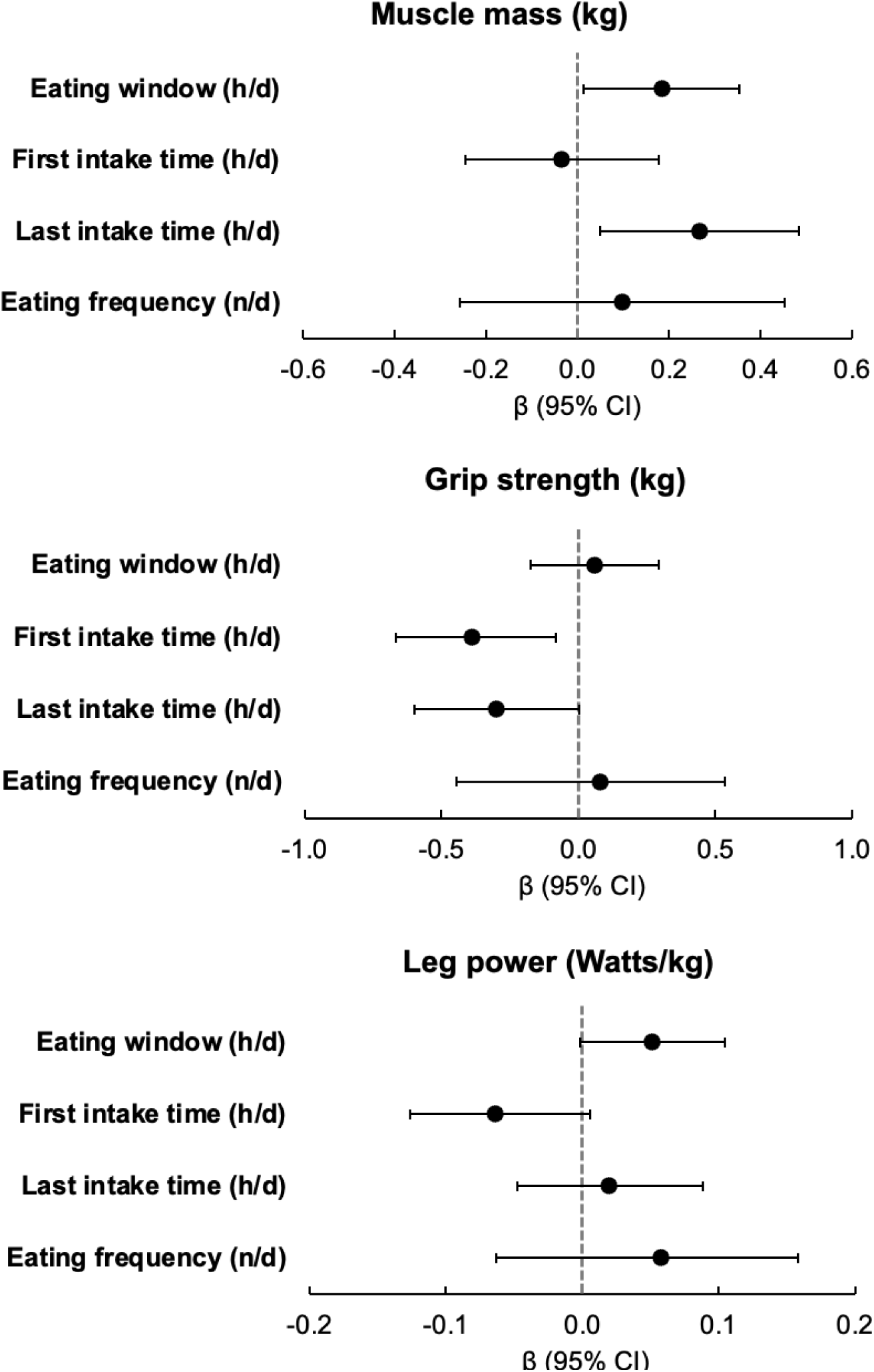
Associations between chrononutrition behaviors, muscle mass, grip strength, and leg power among SOMMA participants. Values are β ± 95% confidence intervals (CI) from multivariable linear regression model; adjusted for age (years), sex (men or women), race (white, black, or others), total daily energy (kcal/d), protein (%kcal/d), and fiber (g/d) intake, weight (kg), height (meters), moderate-to-vigorous physical activity (h/wk), study site (Pittsburgh or Wake Forest), education level (≤ high school and others, some college, college degree, or some graduate/graduate degree), marital status (married, widowed, separated/divorced, or single/never married), and self-reported health status (excellent, very good, good, fair, poor).

**Table 3.** Associations between chrononutrition behaviors and muscle measures among SOMMA participants.

| Chrononutrition behaviors | Models <sup>a</sup> | Muscle mass (kg) |  | Thigh Muscle volume (L) |  | Grip strength (kg) |  | Leg power (Watts/kg) |  |
| --- | --- | --- | --- | --- | --- | --- | --- | --- | --- |
| | | $\beta \pm SE$ | *P* | $\beta \pm SE$ | *P* | $\beta \pm SE$ | *P* | $\beta \pm SE$ | *P* |
| **Eating window (h/d)** | Unadjusted | 0.43 ± 0.12 | <0.001 | 0.17 ± 0.04 | <0.001 | 0.65 ± 0.17 | <0.001 | 0.14 ± 0.03 | <0.001 |
| | Adjusted | 0.18 ± 0.09 | 0.034 | 0.00 ± 0.02 | 0.932 | 0.06 ± 0.12 | 0.628 | 0.05 ± 0.03 | 0.058 |
| **First intake time (h/d)** | Unadjusted | -0.16 ± 0.15 | 0.292 | -0.07 ± 0.05 | 0.186 | -0.77 ± 0.22 | <0.001 | -0.13 ± 0.04 | 0.001 |
| | Adjusted | -0.03 ± 0.11 | 0.746 | 0.01 ± 0.02 | 0.534 | -0.38 ± 0.15 | 0.012 | -0.06 ± 0.03 | 0.075 |
| **Last intake time (h/d)** | Unadjusted | 0.55 ± 0.16 | <0.001 | 0.22 ± 0.05 | <0.001 | 0.32 ± 0.22 | 0.151 | 0.09 ± 0.04 | 0.022 |
| | Adjusted | 0.27 ± 0.11 | 0.017 | 0.02 ± 0.02 | 0.454 | -0.30 ± 0.15 | 0.051 | 0.02 ± 0.03 | 0.557 |
| **Eating frequency (n/d)** | Unadjusted | -0.12 ± 0.25 | 0.618 | 0.06 ± 0.09 | 0.507 | -0.01 ± 0.35 | 0.969 | 0.08 ± 0.06 | 0.183 |
| | Adjusted | 0.10 ± 0.18 | 0.585 | 0.02 ± 0.04 | 0.524 | 0.04 ± 0.25 | 0.860 | 0.05 ± 0.06 | 0.398 |

*Abbreviations:* SE, standard error; SOMMA, Study of Muscle, Morbidity and Aging.
<sup>a</sup> From multivariable linear regression model; adjusted for age (years), sex (men or women), race (white, black, or others), total daily energy (kcal/d), protein (%kcal/d), and fiber (g/d) intake, weight (kg), height (meters), moderate-to-vigorous physical activity (h/wk), study site (Pittsburgh or Wake Forest), education level ( ≤ high school and others, some college, college degree, or some graduate/graduate degree), marital status (married, widowed, separated/divorced, or single/never married), and self-reported health status (excellent, very good, good, fair, poor).

Our analyses revealed a positive association between the time of last food intake and muscle mass in both our unadjusted and fully adjusted regression models (**Table 3** and **Figure 3**). Specifically, we found that each hour later in the time of last food/beverage intake was associated with 0.27 ± 0.11 kg greater muscle mass (*P* = 0.017) after multivariable adjustment. Furthermore, we observed a significant positive association between time of last intake and thigh muscle volume, but this association did not reach statistical significance after controlling for covariates in the multivariable model. Regarding the time of first food/beverage intake and eating frequency, we found no significant associations with muscle mass or thigh muscle volume in either unadjusted or multivariable models.

### Association between chrononutrition behaviors and muscle strength and power

Our unadjusted models showed a positive association between eating window and grip strength (0.65 ± 0.17, *P* < 0.001). However, this association did not remain statistically significant after we controlled for covariates in the multivariable model (**Table 3**). We also observed a significant negative association between the time of first food/beverage intake and maximum grip strength in both unadjusted and multivariable models (−0.38 ± 0.15 kg, *P* = 0.012, **Figure 3**). No significant associations were found between the time of last food/beverage intake or eating frequency and grip strength.

Among our studied chrononutrition behaviors, eating window, as well as the first and last times of food/beverage consumption, showed significant associations with leg power in our unadjusted regression models (**Table 3**). After further adjusting our models for the covariates, only the association between the eating window and leg power remained marginally significant (0.05 ± 0.03 watt/kg, *P* = 0.058). Notably, when adjusting our models for all exercise-related physical activity instead of MVPA, the positive association between the eating window and leg power became significant (data not shown). The associations between the first and last time of food/beverage intake did not remain significant in our multivariable models (**Figure 3**).

## DISCUSSION

Our study revealed that longer eating windows were associated with greater muscle mass and power in community-living older adults. We also found that a later time of the last food/beverage intake was linked to greater muscle mass, while an earlier timing of the first food/beverage intake was associated with higher grip strength. The observed associations between chrononutrition behaviors and muscle health were independent of demographic factors (e.g., age and race), lifestyle (such as total daily energy, protein, and fiber intake and physical activity), and anthropometric measures.

The circadian regulation of skeletal muscle physiology is essential for maintaining optimal muscle structure and function, with the expression of clock genes playing a key role in this process [19]. External cues, such as timing of food/beverage consumption [9], can modulate the temporal expression of these genes, highlighting the potential impact of chrononutrition on muscle circadian rhythms [9, 19]. While the timing of food intake may play a crucial role in maintaining muscle health, most nutritional studies have predominantly concentrated on the quantity and quality of foods to preserve muscle health and prevent sarcopenia in older adults.

### Eating later in the evening and longer eating windows were associated with greater muscle mass in older adults

Here, we performed a comprehensive assessment of various chrononutrition behaviors and their relationships to muscle mass and function in community-dwelling adults. Our study revealed that longer eating window and eating later in the evening were associated with greater muscle mass in older adults. This finding contradicts the reported benefits of intermittent fasting in promoting muscle health which has been mostly supported by non-human studies [23] or studies in younger and physically active adults [24, 25]. Also, most studies on intermittent fasting have focused on its effects on weight and fat loss, with comparatively less attention given to its impact on muscle gain and maintenance. However, the available evidence suggests that intermittent fasting does not significantly differ from non-fasting diets in promoting muscle gain or preventing muscle loss [49]. Also, the eating window of our study participants was, on average, 11 hours per day, which is shorter than the 12-hour daily eating window previously reported by our group among 34,470 US adults (> 19 years) in the National Health and Nutrition Examination Survey (NHANES) [50]. While this suggests that older individuals may have shorter eating windows, further large-scale and representative research is needed to fully elucidate meal timing patterns across different age groups, including young, middle-aged, and older adults.

In this study, eating frequency and the time of first food/beverage intake were not associated with muscle mass. Consistent to our results, it has been reported that increasing the frequency of meals does not have a positive impact on the body composition and fat free mass in sedentary individuals [51, 52]. It is also possible that other factors, such as specific food choices, could play a role in influencing muscle mass regardless of eating frequency or time of first food/beverage intake. Furthermore, our findings showed that the majority of chrononutrition behaviors were significantly associated with muscle mass, as measured by the D_3_Cr method, but not with thigh muscle volume, as measured by MR scans. While these two methods capture different aspects of muscle mass, one potential explanation for the discrepancy in associations could be that D_3_Cr provides a more comprehensive measure of total body muscle mass compared to the localized measurement of thigh muscle volume. Future studies incorporating whole body muscle volume measurements from MR scans could provide a more complete understanding of the relationships between chrononutrition behaviors and muscle mass. On the other hand, it is possible that measurement characteristics of the assessments (e.g., precision and accuracy) depend on chrononutrition habits; the D_3_CR dilution method requires a fasting morning urine sample.

### Earlier first intake times and longer eating windows were associated with better muscle function

There is a wealth of research on meal timing recommendations for elite athletes, aimed at enhancing muscle function and performance [53, 54]. However, the significance of meal timing in preserving and improving muscle function in older adults is not well-understood. In this study, we showed that an earlier first intake time was significantly associated with greater grip strength among older adults; also, a longer eating window was marginally associated with greater leg power among older adults. In contrast to our findings, a recent systematic review showed that time-restricted eating was associated with better physical performance among young athletes [55]. Given the limited human studies examining the impact of time-restricted eating on muscle function in older adults, further research is warranted to investigate the potential benefits of this dietary approach on muscle function in aging. Understanding the relationship between various measures of chrononutrition and muscle function could help inform the development of tailored dietary recommendations aimed at improving muscle function and physical performance in this age group.

The majority of the chrononutrition behaviors investigated here (i.e., eating window, first, and last intake time of food/beverages) were associated with muscle function in our unadjusted regression. However, upon adjusting our models for potential covariates, especially total energy intake, we found that only the eating window and the time of first food intake remained significantly associated with muscle function. Considering that undernutrition, rather than overnutrition, is a primary concern in older adults due to its association with impaired immune and muscle function, delayed wound healing, hospitalization, and mortality [56], adequate intake of energy and nutrients through a well-timed diet may promote muscle health in older adults. However, large-scale multi-center trials are needed to confirm the potential benefits of such interventions. Moreover, to distinguish the associations observed between chrononutrition behaviors and muscle mass and function are independent of the effect of chrononutrition on fat mass, we accounted for body size measures such as weight and height in our analysis. Despite this, the associations between meal timing and muscle remained significant, indicating that the observed effects are not attributed to the presence of obesity.

The impact of food timing on human health and aging is a rapidly emerging area of research. One potential mechanism that is engaged with differently timed feeding patterns and healthy aging is the intrinsic circadian biology system. Studies have shown that consuming small, nutrient-dense foods or beverages, or a single macronutrient (less than 200 kcals) before nighttime sleep, can promote positive physiological changes by providing a bedtime supply of nutrients [57]. For instance, consuming a high-protein beverage 30 minutes before sleep or during sleep (via nasogastric tube) has been demonstrated to increase plasma concentrations of nutrients, particularly amino acids. This, in turn, stimulates and increases overnight muscle protein synthesis, inhibits protein breakdown, aids muscle recovery, and enhances overall metabolism in both young [58] and older men [59]. Although the beneficial effects of certain meal timing patterns, such as fasting diets, have been demonstrated, more research is needed to fully understand the potential of chrono diets across diverse health and disease populations.

### Strengths and limitations

To the best of our knowledge, our study is the first to explore the link between multiple chrononutrition behaviors and muscle mass and function in a large number of older adults. Unlike many cohort studies that collect dietary data using food frequency questionnaires which do not capture the timing of food intake, our study utilized the NIH/NCI developed ASA24 tool. While it is important to note that we only collected two 24-hour dietary recalls, which may not be fully representative of an individual’s usual dietary intake, the use of 24-hour food recall allowed us to obtain precise information on both the quantity and timing of food and beverage consumption, hence enabling a more accurate assessment of chrononutrition patterns. Our study aimed to standardize feeding time to clock time, considering the diversity in the chronotypes which is the tendency of individuals to sleep and be active at particular times of the day, could also impact the meal timing behaviors. Therefore, further research is needed to investigate the optimal timing of food intake for individuals with different chronotypes.

Another strength of our study is the utilization of multiple precise measures to determine muscle mass. We employed MR scans as well as the D_3_Cr dilution method, which is a safe, non-invasive, reliable, and highly accurate method for measuring total body muscle mass. The cross-sectional observational design of the present study limited the ability to assess the causal effect of chrononutrition behaviors on muscle health. Also, our study population primarily consisted of non-Hispanic white older adults. As a result, the generalizability of our findings may be limited, and further research is needed to confirm our results in larger and more diverse populations of older individuals.

## Conclusions

Our findings highlight the importance of meal timing in muscle health in older adults. Here we showed that chrononutrition behaviors, such as longer eating window, earlier time of the first and later time of last food/beverage consumption, were associated with better muscle mass and strength in older adults. Our findings suggest that precision nutrition interventions aimed at optimizing chrononutrition patterns could potentially enhance muscle mass and function, leading to improvements in overall physical performance and health in older adults.

## Data Availability

Data described in the manuscript, code book, and analytic code will be made publicly and freely available without restriction.

## ACKNOWLEDGMENT

The authors’ contributions were as follows - SF: designed the study, ZM and SF: analyzed and wrote the manuscript; PMC, SBK, FGST, KAE, MLE, and ABN: were involved in the interpretation of data and manuscript critical review; SF: had primary responsibility for the final content; and all authors: read and approved the final manuscript.

## The Sources of Support including grants, fellowships

The Study of Muscle, Mobility and Aging is supported by funding from the National Institute on Aging, grant number AG059416.” The following statement should also be included: Study infrastructure support was funded in part by NIA Claude D. Pepper Older American Independence Centers at University of Pittsburgh (P30AG024827) and Wake Forest University (P30AG021332) and the Clinical and Translational Science Institutes, funded by the National Center for Advancing Translational Science, at Wake Forest University (UL1 0TR001420). SF is supported by a career development award from the National Institute on Aging (K01 AG071855) and the Pittsburgh Older Americans Independence Center Scholar (P30AG024827; Sub# 6306]).

## Disclaimers

No conflict of interest.

## Abbreviation list and their definitions

ASA24: Self-administered 24-hour Food Recall
BMI: Body mass index
BW: Body weight
CI: Confidence interval
CESD: Center for Epidemiologic Studies Depression Scale
CHAMPS: Community Healthy Activities Model Program for Seniors
D_3_Cr: D_3_-creatine
HPLC: High-performance liquid chromatography
IGF-1: Insulin-like growth factor 1
MR: Magnetic resonance imaging
MS: Mass spectrometry
MVPA: Moderate-to-vigorous physical activity
SD: Standard deviation
SE: Standard error
SOMMA: Study of Muscle, Mobility, and Aging
USDA: United States Department of Agriculture
1-RM: 1 repetition maximum

---

**eFigure 1.**
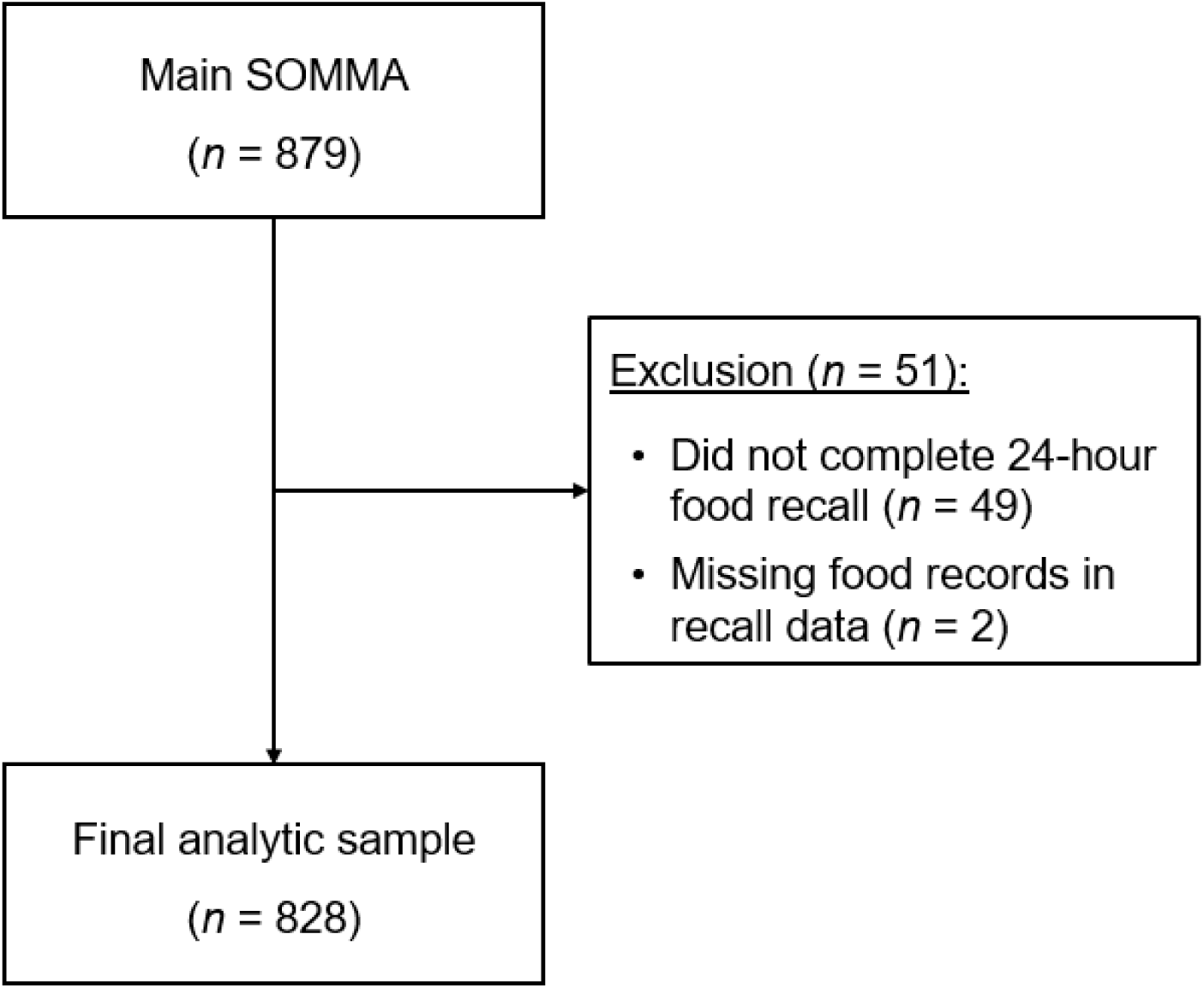
Flowchart depicting the exclusion and inclusion of SOMMA study participants in the analysis. SOMMA, Study of Muscle, Mobility, and Aging.

